# Cardio-Renal Anemia Syndrome in Patients with Atrial Functional Mitral Regurgitation: Subanalysis from the REVEAL-AFMR Registry

**DOI:** 10.1101/2025.08.11.25333466

**Authors:** Madoka Sano, Taiji Okada, Tomohiro Kaneko, Masashi Amano, Yukio Sato, Yohei Ono, Masaru Obokata, Kimi Sato, Kojiro Morita, Tomoko Machino, Yukio Abe, Yutaka Furukawa, Nobuyuki Kagiyama, the REVEAL-AFMR study group

## Abstract

**Background:** Atrial functional mitral regurgitation (AFMR) frequently affects older patients with heart failure (HF), causing venous congestion and reduced forward cardiac output. This hemodynamic compromise may accelerate chronic kidney disease (CKD) and impair oxygen delivery, thereby exacerbating anemia. Cardio-renal anemia syndrome (CRAS) is a prognostic factor in HF, and specific hemodynamic disturbances in AFMR may amplify its consequences. In this study, we aimed to evaluate the impact of CRAS on cardiovascular outcomes in patients with AFMR.

**Methods:** The Real-World Observational Study for Investigating the Prevalence and Therapeutic Options for Atrial Functional Mitral Regurgitation (REVEAL-AFMR) registry is a multicenter, retrospective, observational cohort that enrolled patients with moderate-to-severe AFMR in 2019. Patients were stratified into the CRAS and non-CRAS groups based on the simultaneous presence of HF, CKD, and anemia. The primary outcome was a composite of all-cause mortality and hospitalization for HF.

**Results:** Among the 986 patients analyzed, 258 had CRAS. Over a median follow-up period of 1,040 days, 327 composite events occurred. Kaplan–Meier analysis demonstrated a higher incidence of composite cardiovascular outcomes in patients with CRAS than in those without CRAS, consistent across all severities of AFMR. In multivariate Cox proportional hazards regression analysis, CRAS was independently associated with cardiovascular events, along with older age, lower body mass index, and lower systolic blood pressure.

**Conclusion:** CRAS is common in patients with AFMR and may be independently associated with an increased risk of future cardiovascular events. Assessment of CRAS may be helpful for risk stratification and management in this population.

## Introduction

The incidence of heart failure (HF), atrial fibrillation, and valvular heart disease is increasing in aging societies. Atrial functional mitral regurgitation (AFMR) is a common complication in patients with HF, and previous studies have suggested its association with worse clinical outcomes, including HF hospitalization, cardiovascular death, and all-cause mortality (1). Patients with AFMR are typically older and have multiple comorbidities that compromise prognosis. Among these comorbidities, chronic kidney disease (CKD) and anemia frequently coexist, and each has been independently associated with adverse cardiovascular events. CKD is a well-known risk factor for HF progression and is associated with poorer outcomes in other cardiovascular diseases (2). Anemia, caused by iron deficiency, inflammation, renal insufficiency, and other factors, reduces oxygen delivery, increases cardiac oxygen demand, and promotes left ventricular remodeling, thereby worsening HF symptoms and prognosis (3).

The coexistence of HF, CKD, and anemia may have a synergistic effect, amplifying the risk of cardiovascular events—a condition termed cardio-renal anemia syndrome (CRAS) (4,5). Although CRAS strongly predicts adverse cardiovascular events in various populations with HF, its specific impact on patients with AFMR remains unclear. In patients with AFMR, chronic left atrial (LA) pressure elevation and volume overload impose backward pressure on the pulmonary and renal venous systems, leading to renal venous congestion and reduced effective renal perfusion (6). Even in the absence of overt left ventricular failure, these changes accelerate CKD progression and contribute to the development and worsening of anemia. AFMR also reduces forward cardiac output and increases the cardiac workload, further compromising oxygen delivery. Therefore, anemia may exert a disproportionately negative impact in this setting. These pathophysiological disturbances suggest that CRAS may exert particularly harmful effects in patients with AFMR. Considering these compounded pathophysiological mechanisms, clarifying the prognostic value of CRAS in patients with AFMR is important.

Therefore, we aimed to analyze a large AFMR cohort to determine the prevalence of CRAS and examine its combined impact on cardiovascular outcomes, to identify modifiable prognostic factors.

## Methods

### Study design

This was a secondary analysis of the Real-World Observational Study for Investigating the Prevalence and Therapeutic Options for Atrial Functional Mitral Regurgitation (REVEAL-AFMR) registry. The details of the main study design and primary results have been published previously (7). In brief, it was a multicenter, retrospective, observational cohort study conducted in Japan at 26 participating institutions. It included patients aged ≥ 20 years with moderate or severe AFMR, defined by preserved left ventricular ejection fraction (≥ 50%) without regional wall motion abnormality, LA dilatation (LA volume index ≥ 38 mL/m^2^ for men, ≥ 41 mL/m^2^ for women, based on reported normal values in the Japanese population) (8), and absence of degenerative valvular changes, between January 2019 and December 2019. The exclusion criteria were as follows: trivial-to-mild mitral regurgitation (MR); degenerative MR or post-mitral valve surgery; absence of LA dilatation; other etiologies, including systolic anterior motion; or acute decompensated HF. Patients without hemoglobin and/or estimated glomerular filtration rate (eGFR) data were also excluded.

Patients were stratified into two groups based on whether they met the criteria for CRAS, defined as the simultaneous presence of HF, CKD, and anemia. HF was defined as New York Heart Association (NYHA) class II or higher, or a documented history of HF (9). CKD was defined as eGFR < 60 mL/min/1.73 m^2^, calculated using the CKD-EPI Creatinine Equation (10). Anemia was defined as hemoglobin < 13 g/dL in men and < 12 g/dL in women (11).

The primary outcome was a composite of all-cause death (ACD) and hospitalization for HF. The secondary outcomes were ACD and hospitalization for HF, analyzed separately.

This study was registered in the University Hospital Medical Information Network Clinical Trials Registry and the Japanese Clinical Trial Registry (registration number: UMIN000046146) and was approved by the Institutional Review Board of Juntendo University and each participating institution, including Kobe City Medical Center General Hospital (approval number: kzh250408). The investigation conformed to the principles outlined in the Declaration of Helsinki (Br Med J 1964; ii:177) and adhered to the Strengthening the Reporting of Observational Studies in Epidemiology reporting guidelines. All enrolled individuals were informed of the study details and reassured that they could withdraw at any time. Written informed consent was waived because of the observational and retrospective nature of this study, in accordance with the ethical guidelines for medical and biological research involving human participants in Japan.

### Statistical analyses

Categorical variables are expressed as numbers and percentages and compared using the chi-squared or Fisher’s exact test, as appropriate. Continuous variables are expressed as medians with interquartile ranges (IQRs) or means with standard deviations and were compared using the Wilcoxon rank-sum test or Student’s t-test, depending on their distribution. Cumulative incidences were estimated using the Kaplan–Meier method, and differences among groups were assessed using a log-rank test. Multivariate Cox proportional hazards models were used to assess the primary and secondary outcomes. The multivariate analysis was adjusted for age, sex, CRAS, body mass index (BMI), systolic blood pressure, and severity of mitral regurgitation (MR), based on baseline data at the time of cohort entry, as used in previous analyses of the REVEAL-AFMR registry (7). To evaluate the combined and interactive effects of HF, CKD, and anemia, additional Cox models were used to examine each component and their interactions, testing whether CRAS represented a synergistic risk factor beyond its individual components. All p-values were two-sided, with <0.05 indicating statistical significance. Missing values were not imputed and were excluded from the analysis. All statistical analyses were performed using JMP software (version 18.0, SAS Institute Japan. Tokyo, Japan).

## Results

### Characteristics of the study population

Figure 1 shows the patient selection flowchart. Of the 177,235 patients who underwent transthoracic echocardiography, 8,867 had moderate or severe MR, and 1,007 were diagnosed with AFMR. After excluding 21 patients with missing hemoglobin or eGFR data, 986 patients were included in the analysis. Of the 986 patients, 724 (73.4%) had HF, 429 (43.5%) had CKD, and 570 (57.8%) had anemia (see Central Illustration). CRAS, defined as the coexistence of all three conditions, was present in 258 (26.2%) patients. The median follow-up period was 1,040 (IQR, 715–1,173) days.

**Figure 1.**
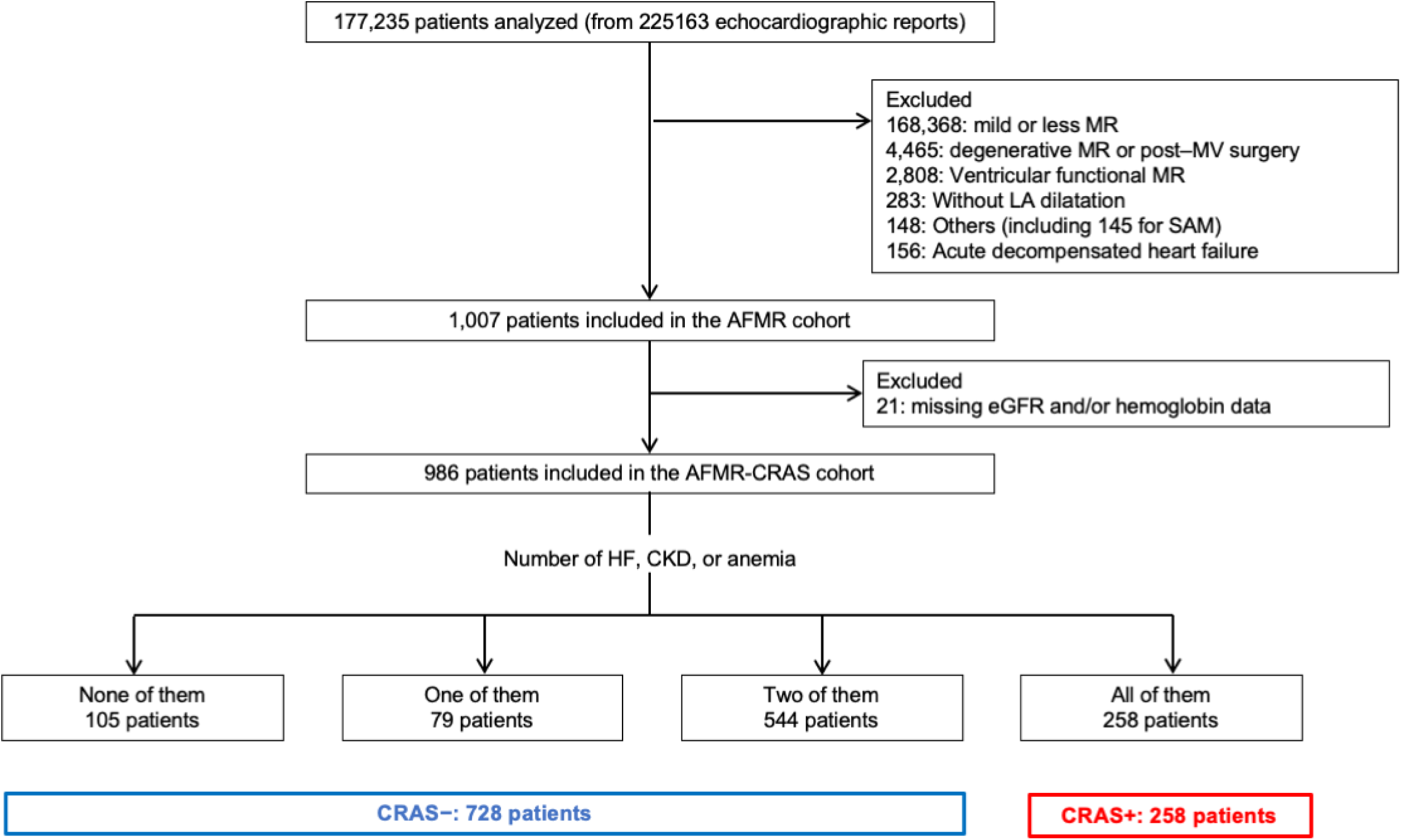
Patients’ flowchart. The REVEAL-AFMR registry enrolled patients with moderate-to-severe AFMR. A total of 986 patients were included in the analysis, of whom 258 (26.2%) were diagnosed with CRAS. REVEAL-AFMR, Real-World Observational Study for Investigating the Prevalence and Therapeutic Options for Atrial Functional Mitral Regurgitation; AFMR, atrial functional mitral regurgitation; MR, mitral regurgitation; MV, mitral valve; LA, left atrium; SAM, systolic anterior motion; CRAS, cardio-renal anemia syndrome; HF, heart failure; CKD, chronic kidney disease.

**Central Illustration.**
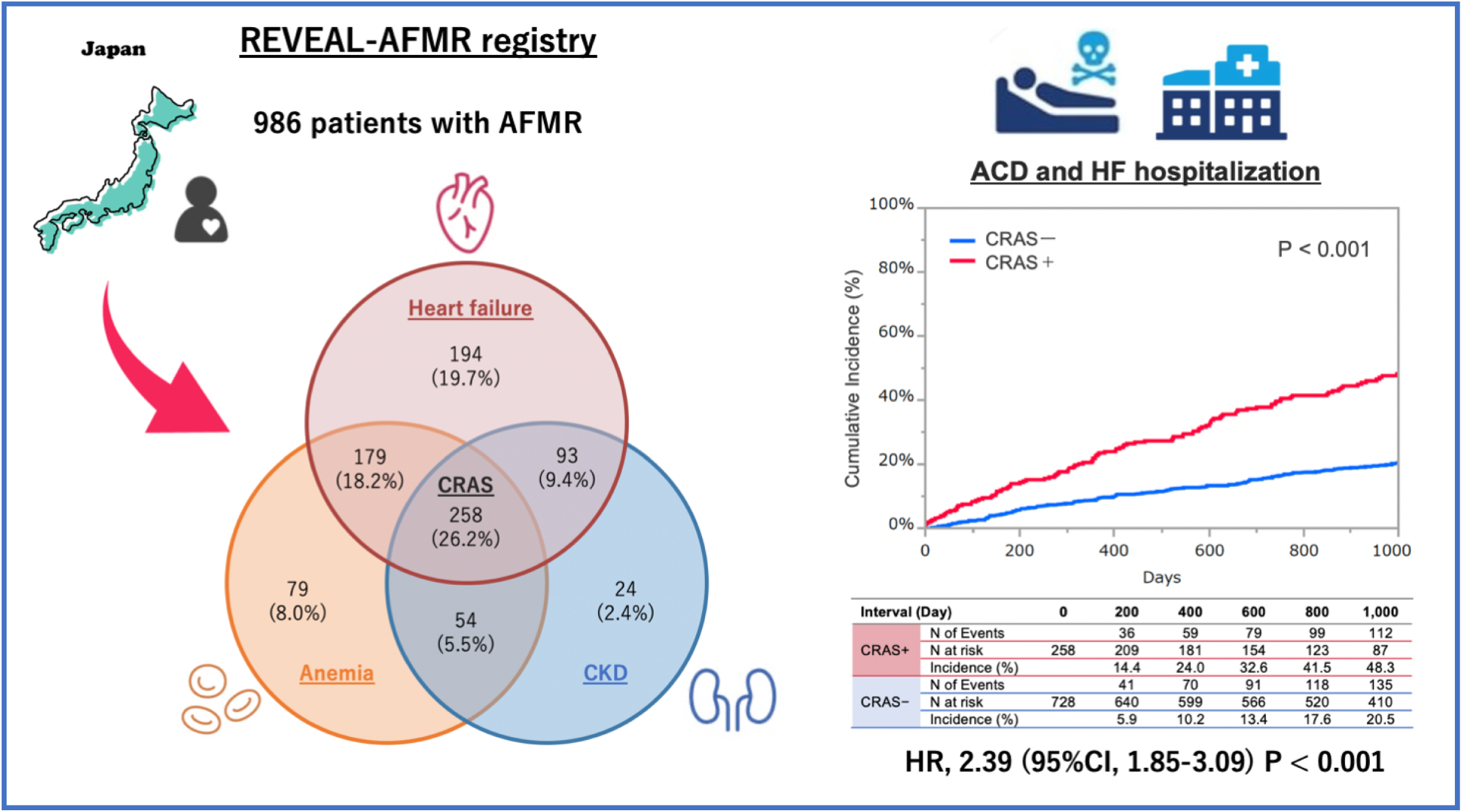
The REVEAL-AFMR registry is an observational, multicenter, retrospective registry from January 2019 to December 2019, that enrolled patients with moderate to severe atrial functional mitral regurgitation (AFMR). Cardio-renal anemia syndrome was present in 26.2% of patients with AFMR and was significantly associated with worse cardiovascular outcomes (hazard ratio 2.39, 95% confidence interval 1.85-3.09, p < 0.001). AFMR, atrial functional mitral regurgitation; CKD, chronic kidney disease; CRAS, cardiorenal anemia syndrome; ACD, all-cause death; HF, heart failure; HR, hazard ratio; CI, confidence interval.

Patients’ baseline characteristics, classified based on the presence of CRAS, are presented in Table 1. Patient characteristics classified into four groups according to the number of CRAS components are presented in Supplemental Table 1. The mean age was 77.7 ± 9.5 years, and 438 (44.4%) patients were men. Compared with patients without CRAS, those with CRAS were older and had a higher prevalence of NYHA III or IV, pacemaker implantation, coronary artery disease, dementia, dialysis, chronic obstructive pulmonary disease, and gastrointestinal bleeding. On echocardiography, patients with CRAS had a larger LA volume index and a higher prevalence of severe MR and tricuspid regurgitation compared with those without CRAS. Diuretics were used more frequently in patients with CRAS than in those without.

**Table 1.**
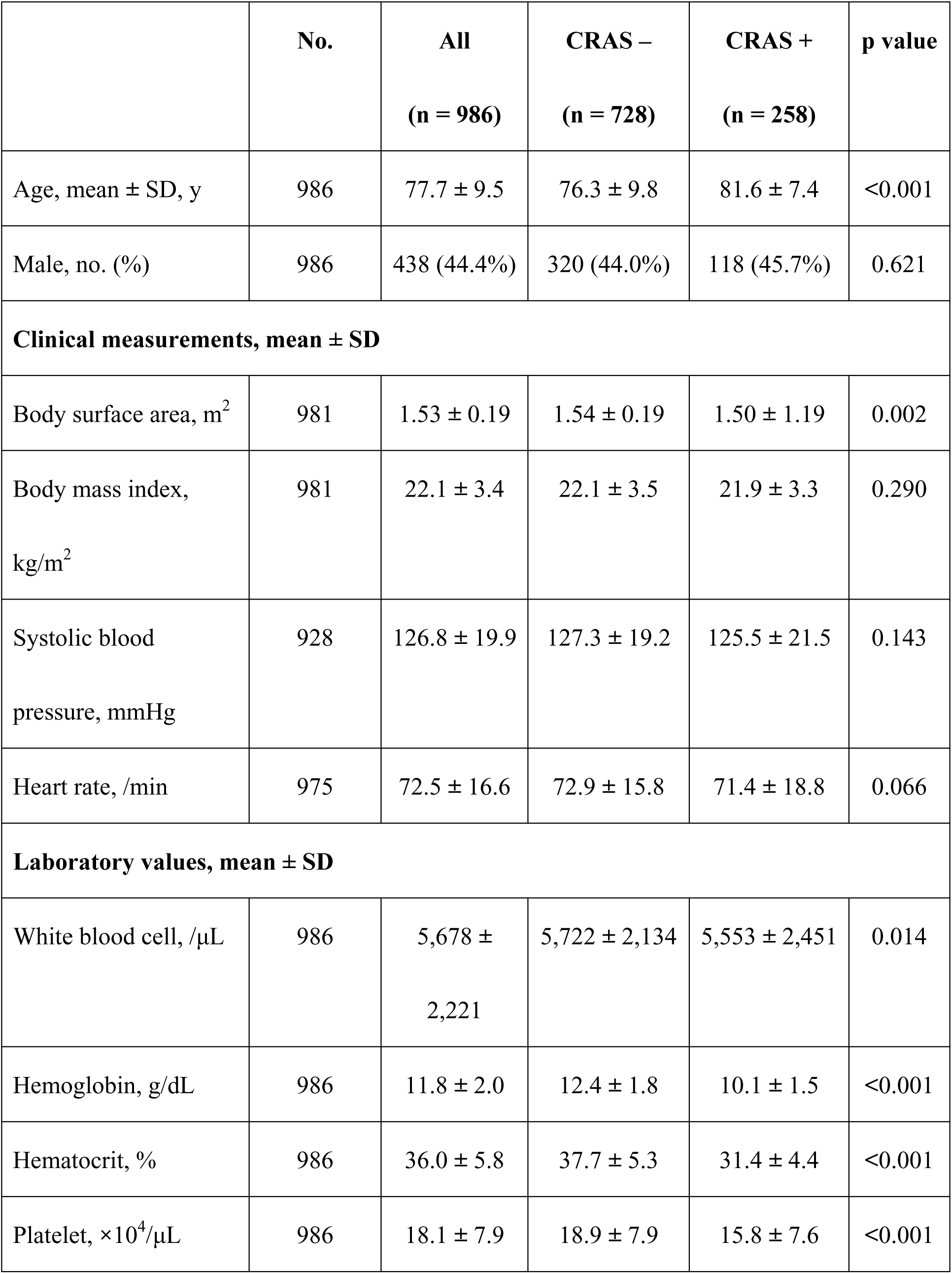
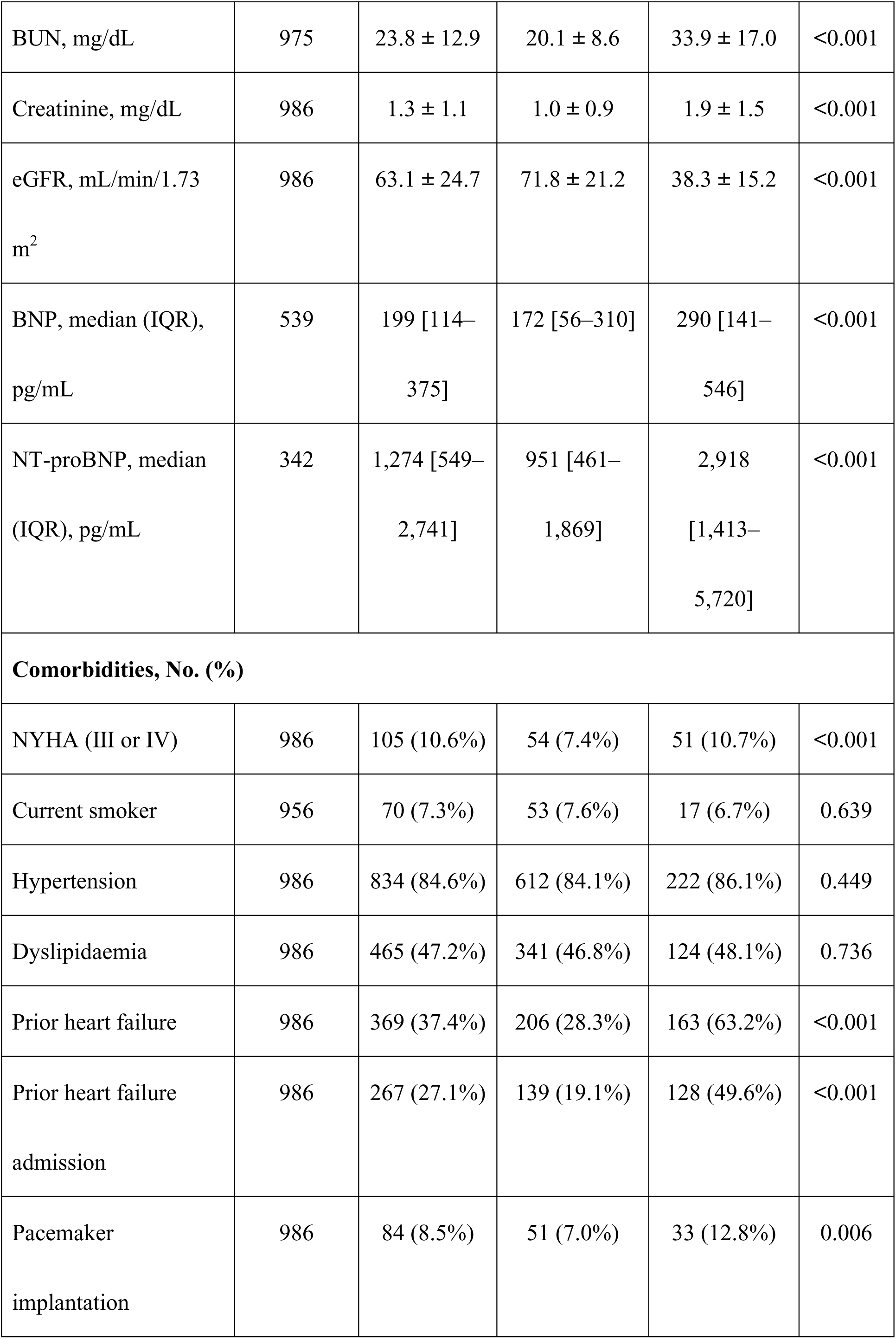
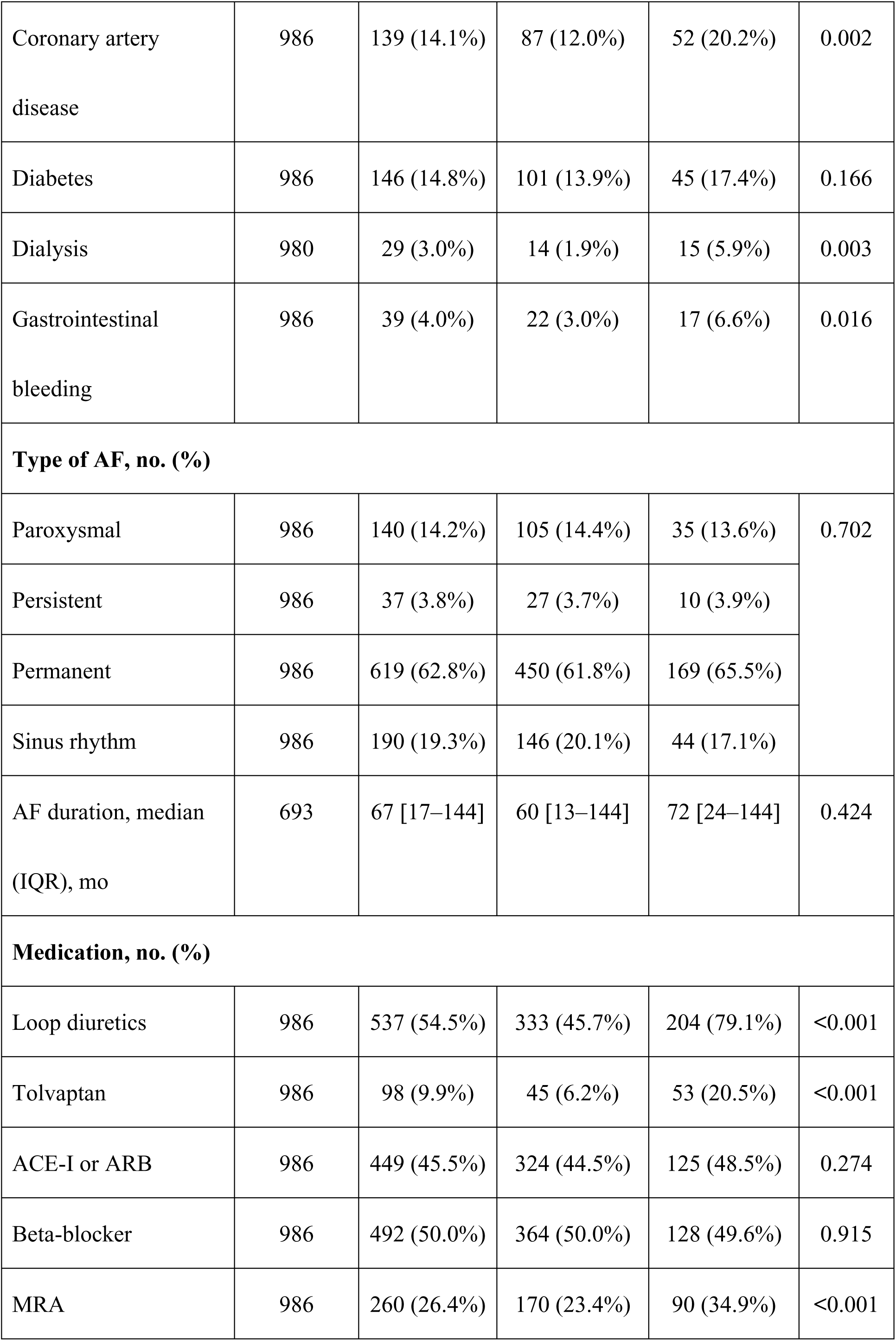
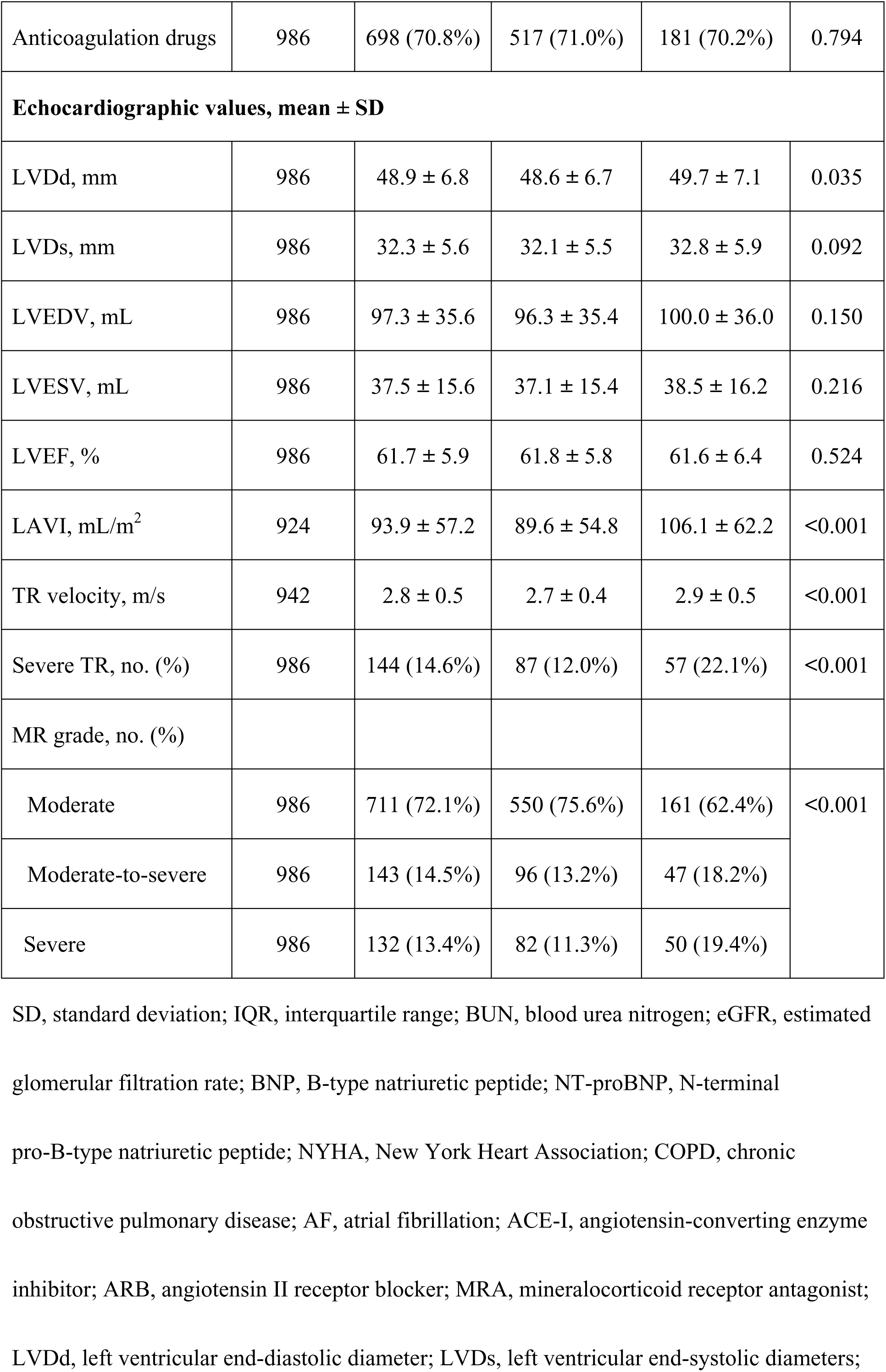
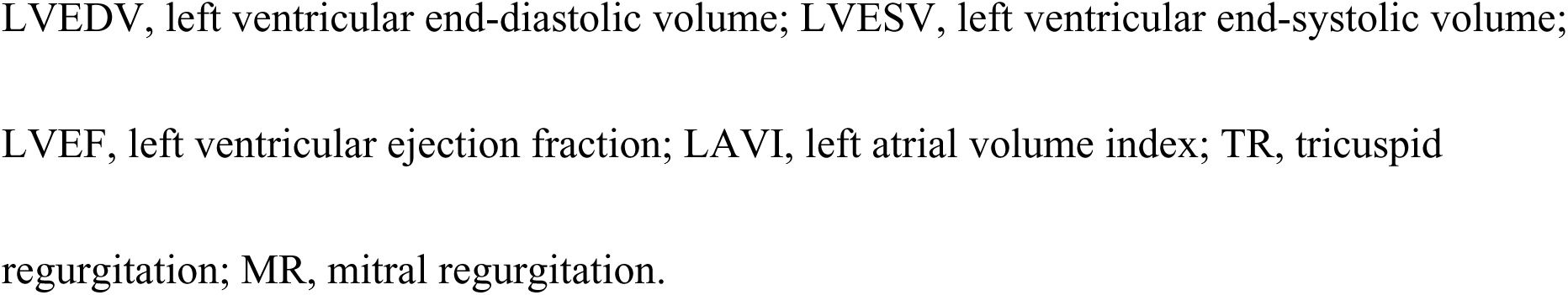
Patient characteristics in each stratum.

|  | No. | All<br>(n = 986) | CRAS –<br>(n = 728) | CRAS +<br>(n = 258) | p value |
| --- | --- | --- | --- | --- | --- |
| Age, mean ± SD, y | 986 | 77.7 ± 9.5 | 76.3 ± 9.8 | 81.6 ± 7.4 | <0.001 |
| Male, no. (%) | 986 | 438 (44.4%) | 320 (44.0%) | 118 (45.7%) | 0.621 |
| **Clinical measurements, mean ± SD** |  |  |  |  |  |
| Body surface area, m<sup>2</sup> | 981 | 1.53 ± 0.19 | 1.54 ± 0.19 | 1.50 ± 1.19 | 0.002 |
| Body mass index,<br>kg/m<sup>2</sup> | 981 | 22.1 ± 3.4 | 22.1 ± 3.5 | 21.9 ± 3.3 | 0.290 |
| Systolic blood<br>pressure, mmHg | 928 | 126.8 ± 19.9 | 127.3 ± 19.2 | 125.5 ± 21.5 | 0.143 |
| Heart rate, /min | 975 | 72.5 ± 16.6 | 72.9 ± 15.8 | 71.4 ± 18.8 | 0.066 |
| **Laboratory values, mean ± SD** |  |  |  |  |  |
| White blood cell, /μL | 986 | 5,678 ±<br>2,221 | 5,722 ± 2,134 | 5,553 ± 2,451 | 0.014 |
| Hemoglobin, g/dL | 986 | 11.8 ± 2.0 | 12.4 ± 1.8 | 10.1 ± 1.5 | <0.001 |
| Hematocrit, % | 986 | 36.0 ± 5.8 | 37.7 ± 5.3 | 31.4 ± 4.4 | <0.001 |
| Platelet, $\times 10^4$/μL | 986 | 18.1 ± 7.9 | 18.9 ± 7.9 | 15.8 ± 7.6 | <0.001 |
| BUN, mg/dL | 975 | 23.8 ± 12.9 | 20.1 ± 8.6 | 33.9 ± 17.0 | <0.001 |
| Creatinine, mg/dL | 986 | 1.3 ± 1.1 | 1.0 ± 0.9 | 1.9 ± 1.5 | <0.001 |
| eGFR, mL/min/1.73<br>m<sup>2</sup> | 986 | 63.1 ± 24.7 | 71.8 ± 21.2 | 38.3 ± 15.2 | <0.001 |
| BNP, median (IQR),<br>pg/mL | 539 | 199 [114–<br>375] | 172 [56–310] | 290 [141–<br>546] | <0.001 |
| NT-proBNP, median<br>(IQR), pg/mL | 342 | 1,274 [549–<br>2,741] | 951 [461–<br>1,869] | 2,918<br>[1,413–<br>5,720] | <0.001 |
| **Comorbidities, No. (%)** |  |  |  |  |  |
| NYHA (III or IV) | 986 | 105 (10.6%) | 54 (7.4%) | 51 (10.7%) | <0.001 |
| Current smoker | 956 | 70 (7.3%) | 53 (7.6%) | 17 (6.7%) | 0.639 |
| Hypertension | 986 | 834 (84.6%) | 612 (84.1%) | 222 (86.1%) | 0.449 |
| Dyslipidaemia | 986 | 465 (47.2%) | 341 (46.8%) | 124 (48.1%) | 0.736 |
| Prior heart failure | 986 | 369 (37.4%) | 206 (28.3%) | 163 (63.2%) | <0.001 |
| Prior heart failure<br>admission | 986 | 267 (27.1%) | 139 (19.1%) | 128 (49.6%) | <0.001 |
| Pacemaker<br>implantation | 986 | 84 (8.5%) | 51 (7.0%) | 33 (12.8%) | 0.006 |
| Coronary artery disease | 986 | 139 (14.1%) | 87 (12.0%) | 52 (20.2%) | 0.002 |
| Diabetes | 986 | 146 (14.8%) | 101 (13.9%) | 45 (17.4%) | 0.166 |
| Dialysis | 980 | 29 (3.0%) | 14 (1.9%) | 15 (5.9%) | 0.003 |
| Gastrointestinal bleeding | 986 | 39 (4.0%) | 22 (3.0%) | 17 (6.6%) | 0.016 |
| **Type of AF, no. (%)** |  |  |  |  |  |
| Paroxysmal | 986 | 140 (14.2%) | 105 (14.4%) | 35 (13.6%) | 0.702 |
| Persistent | 986 | 37 (3.8%) | 27 (3.7%) | 10 (3.9%) |  |
| Permanent | 986 | 619 (62.8%) | 450 (61.8%) | 169 (65.5%) |  |
| Sinus rhythm | 986 | 190 (19.3%) | 146 (20.1%) | 44 (17.1%) |  |
| AF duration, median (IQR), mo | 693 | 67 [17–144] | 60 [13–144] | 72 [24–144] | 0.424 |
| **Medication, no. (%)** |  |  |  |  |  |
| Loop diuretics | 986 | 537 (54.5%) | 333 (45.7%) | 204 (79.1%) | <0.001 |
| Tolvaptan | 986 | 98 (9.9%) | 45 (6.2%) | 53 (20.5%) | <0.001 |
| ACE-I or ARB | 986 | 449 (45.5%) | 324 (44.5%) | 125 (48.5%) | 0.274 |
| Beta-blocker | 986 | 492 (50.0%) | 364 (50.0%) | 128 (49.6%) | 0.915 |
| MRA | 986 | 260 (26.4%) | 170 (23.4%) | 90 (34.9%) | <0.001 |
| Anticoagulation drugs | 986 | 698 (70.8%) | 517 (71.0%) | 181 (70.2%) | 0.794 |
| **Echocardiographic values, mean ± SD** |  |  |  |  |  |
| LVDd, mm | 986 | 48.9 ± 6.8 | 48.6 ± 6.7 | 49.7 ± 7.1 | 0.035 |
| LVDs, mm | 986 | 32.3 ± 5.6 | 32.1 ± 5.5 | 32.8 ± 5.9 | 0.092 |
| LVEDV, mL | 986 | 97.3 ± 35.6 | 96.3 ± 35.4 | 100.0 ± 36.0 | 0.150 |
| LVESV, mL | 986 | 37.5 ± 15.6 | 37.1 ± 15.4 | 38.5 ± 16.2 | 0.216 |
| LVEF, % | 986 | 61.7 ± 5.9 | 61.8 ± 5.8 | 61.6 ± 6.4 | 0.524 |
| LAVI, mL/m<sup>2</sup> | 924 | 93.9 ± 57.2 | 89.6 ± 54.8 | 106.1 ± 62.2 | <0.001 |
| TR velocity, m/s | 942 | 2.8 ± 0.5 | 2.7 ± 0.4 | 2.9 ± 0.5 | <0.001 |
| Severe TR, no. (%) | 986 | 144 (14.6%) | 87 (12.0%) | 57 (22.1%) | <0.001 |
| MR grade, no. (%) |  |  |  |  |  |
| Moderate | 986 | 711 (72.1%) | 550 (75.6%) | 161 (62.4%) | <0.001 |
| Moderate-to-severe | 986 | 143 (14.5%) | 96 (13.2%) | 47 (18.2%) |  |
| Severe | 986 | 132 (13.4%) | 82 (11.3%) | 50 (19.4%) |  |

SD, standard deviation; IQR, interquartile range; BUN, blood urea nitrogen; eGFR, estimated glomerular filtration rate; BNP, B-type natriuretic peptide; NT-proBNP, N-terminal pro-B-type natriuretic peptide; NYHA, New York Heart Association; COPD, chronic obstructive pulmonary disease; AF, atrial fibrillation; ACE-I, angiotensin-converting enzyme inhibitor; ARB, angiotensin II receptor blocker; MRA, mineralocorticoid receptor antagonist; LVDd, left ventricular end-diastolic diameter; LVDs, left ventricular end-systolic diameters;
LVEDV, left ventricular end-diastolic volume; LVESV, left ventricular end-systolic volume; LVEF, left ventricular ejection fraction; LAVI, left atrial volume index; TR, tricuspid regurgitation; MR, mitral regurgitation.

### Association between CRAS and cardiovascular outcomes

The Kaplan–Meier curve demonstrated that the cumulative incidence rate of composite cardiovascular events was higher in patients with CRAS than in those without CRAS (1-year event rates: 23.1% vs. 9.3%; log-rank, p < 0.001) (Figure 2). Separately, ACD (1-year event rate: 14.2% vs. 5.4%; log-rank, p < 0.001) and HF hospitalization (1-year event rate, 12.9% vs. 4.5%; log-rank, p < 0.001) were more frequent in patients with CRAS than in those without. Furthermore, stratification according to the number of CRAS components revealed that the coexistence of all three components was associated with a significantly higher incidence of cardiovascular events (log-rank test, p < 0.001) (Supplemental Figure 1). Similarly, individual evaluation of HF, CKD, and anemia showed that each condition was associated with increased composite event incidence (Supplemental Figure 2). Analysis according to MR severity revealed similar CRAS-related risks across the moderate (n = 711, 72.1%), moderate-to-severe (n = 143, 14.5%), and severe MR (n = 132, 13.4%) subgroups (Figure 3A). Similarly, CRAS predicted higher event rates regardless of treatment strategy: medical therapy (n = 845, 85.7%), surgical repair (n = 111, 11.3%), or transcatheter mitral valve repair (n = 30, 3.0%) (Figure 3B).

**Figure 2.**
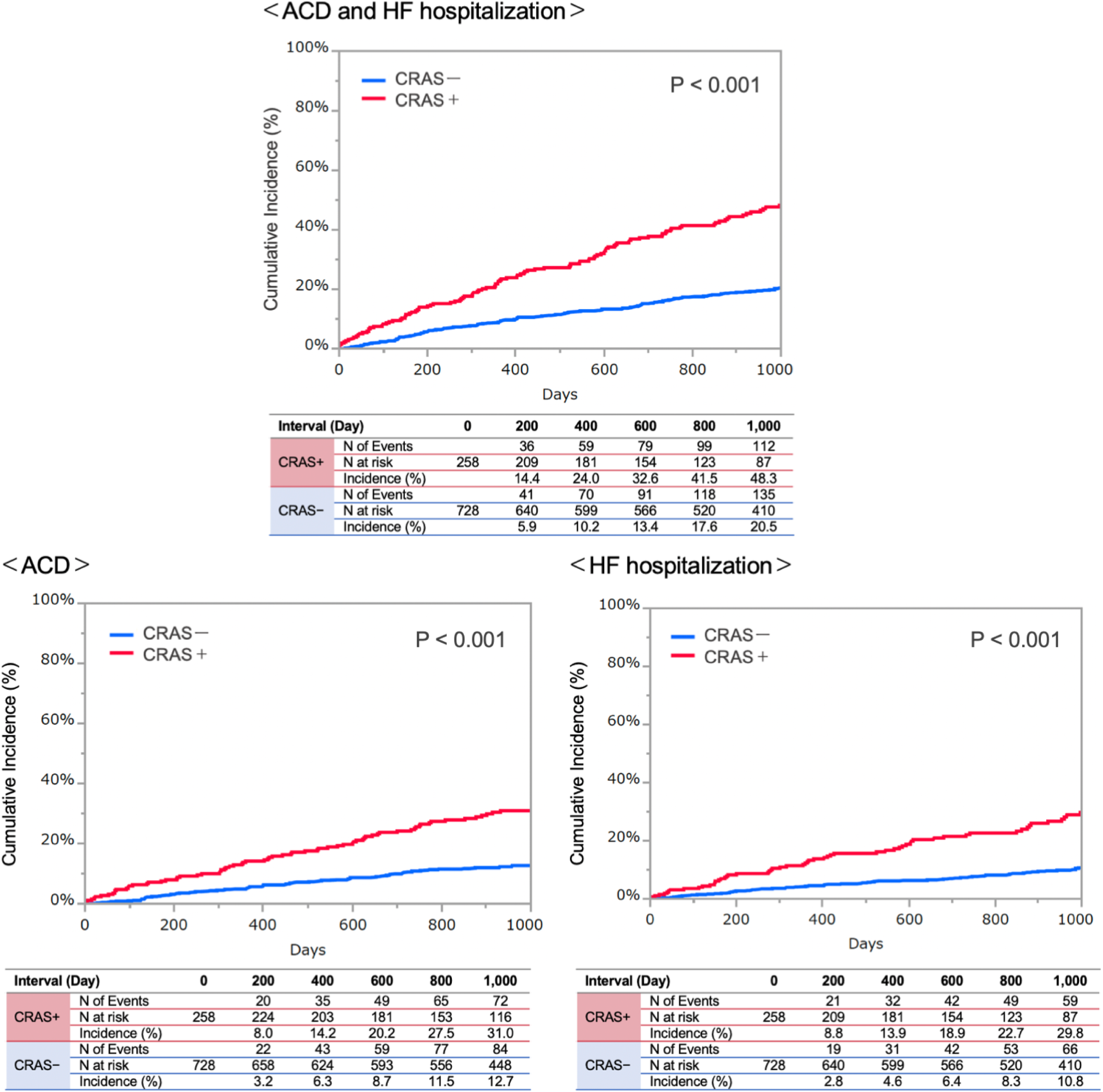
Cumulative incidences of cardiovascular events in patients with/without CRAS. The cumulative incidence of cardiovascular events was higher in patients with CRAS than in those without. CRAS, cardio-renal anemia syndrome; ACD, all-cause death; HF, heart failure.

**Figure 3.**
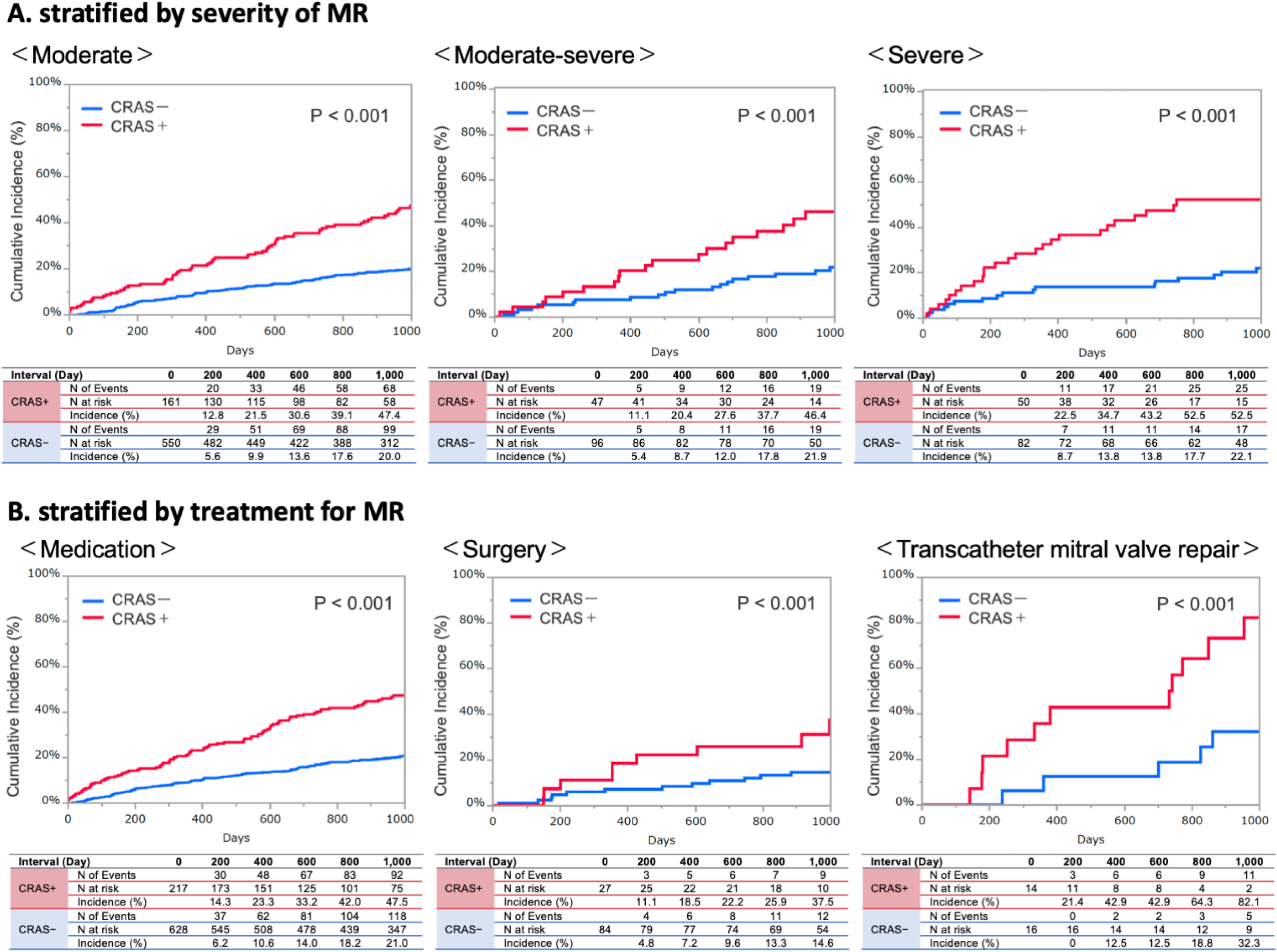
Cumulative incidences of cardiovascular events in patients with/without CRAS stratified by MR severity and treatment. The cumulative incidence of cardiovascular events was higher in patients with CRAS across all degrees of MR severity than in those without, regardless of whether the patients underwent medical therapy, transcatheter mitral valve repair, or mitral valve surgery. CRAS, cardio-renal anemia syndrome; MR, mitral regurgitation.

In multivariate Cox proportional hazards models, CRAS was an independent predictor of poorer outcomes, including composite cardiovascular events (hazard ratio [HR], 2.24; 95% confidence interval [CI], 1.73–2.90; p < 0.001), ACD (HR, 2.14; 95% CI, 1.56–2.94; p < 0.001), and HF hospitalization (HR, 1.38; 95% CI, 1.07–1.79; p = 0.010) (Table 2). Other independent predictors of composite cardiovascular events included older age, lower BMI, and lower systolic blood pressure.

**Table 2:**
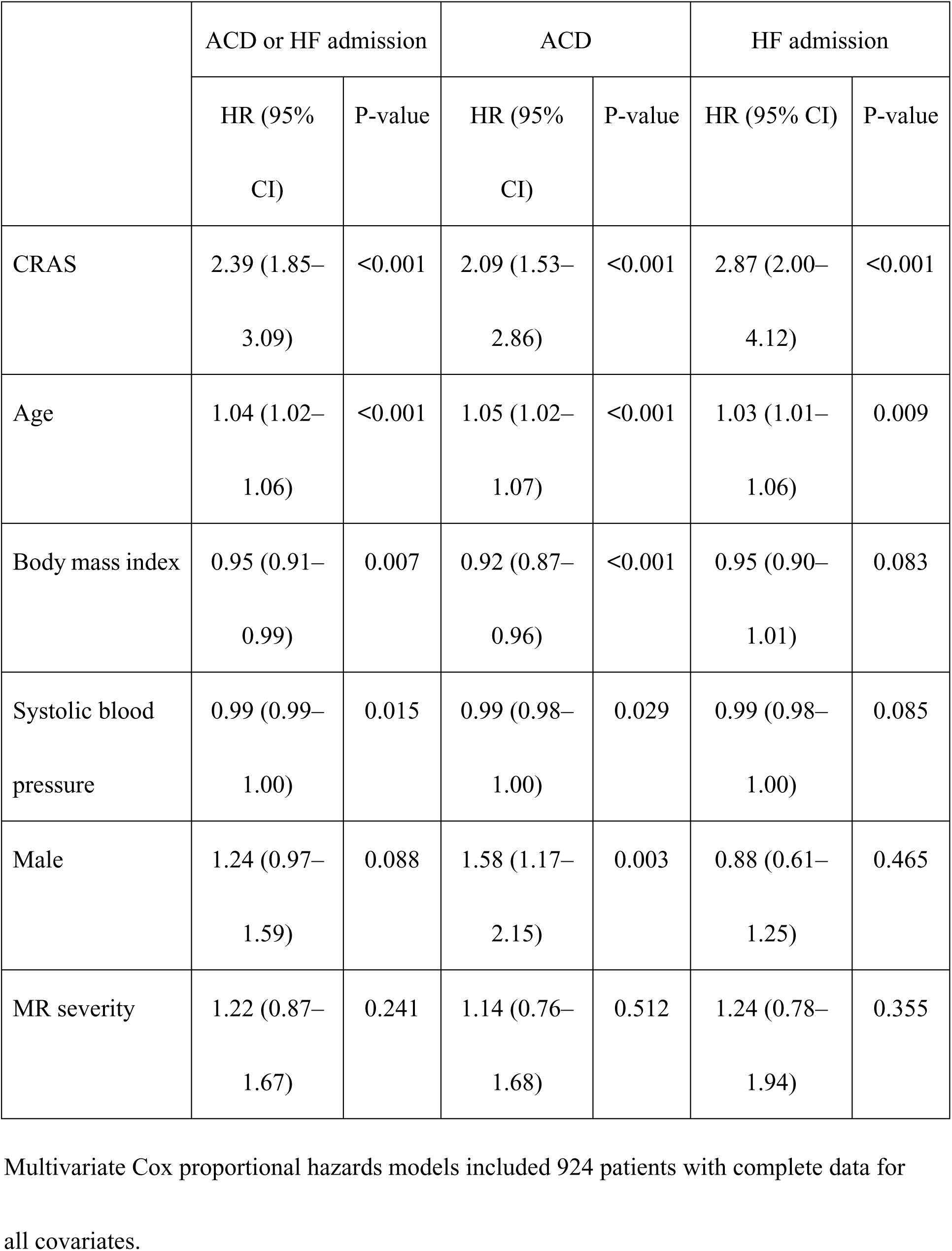
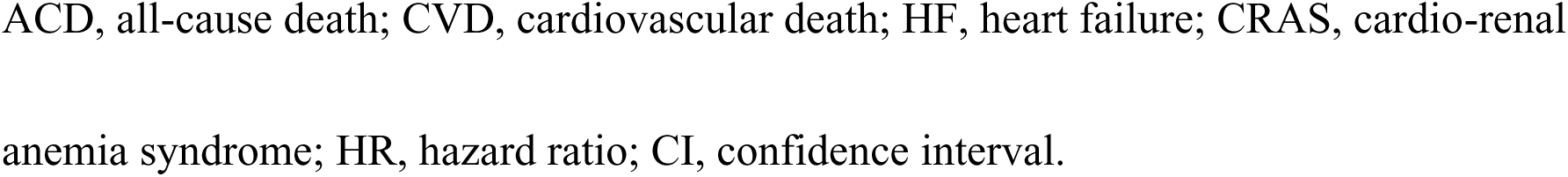
Potential risk factors for all-cause death and heart failure admission.

|  | ACD or HF admission |  | ACD |  | HF admission |  |
| --- | --- | --- | --- | --- | --- | --- |
|  | HR (95% CI) | P-value | HR (95% CI) | P-value | HR (95% CI) | P-value |
| CRAS | 2.39 (1.85–3.09) | <0.001 | 2.09 (1.53–2.86) | <0.001 | 2.87 (2.00–4.12) | <0.001 |
| Age | 1.04 (1.02–1.06) | <0.001 | 1.05 (1.02–1.07) | <0.001 | 1.03 (1.01–1.06) | 0.009 |
| Body mass index | 0.95 (0.91–0.99) | 0.007 | 0.92 (0.87–0.96) | <0.001 | 0.95 (0.90–1.01) | 0.083 |
| Systolic blood pressure | 0.99 (0.99–1.00) | 0.015 | 0.99 (0.98–1.00) | 0.029 | 0.99 (0.98–1.00) | 0.085 |
| Male | 1.24 (0.97–1.59) | 0.088 | 1.58 (1.17–2.15) | 0.003 | 0.88 (0.61–1.25) | 0.465 |
| MR severity | 1.22 (0.87–1.67) | 0.241 | 1.14 (0.76–1.68) | 0.512 | 1.24 (0.78–1.94) | 0.355 |

Multivariate Cox proportional hazards models included 924 patients with complete data for
all covariates.
ACD, all-cause death; CVD, cardiovascular death; HF, heart failure; CRAS, cardio-renal
anemia syndrome; HR, hazard ratio; CI, confidence interval.

### Interaction of CRAS components

Interaction analysis of CRAS components showed that HF (HR, 2.29; 95% CI, 1.11– 5.32; p = 0.020) and anemia (HR, 2.39; 95% CI, 1.00–6.04; p = 0.050) were independently associated with poorer outcomes, whereas CKD alone was not statistically significance (HR, 1.97; 95% CI, 0.52–6.29; p = 0.290) (Table 3). None of the interaction terms for the two-component combinations (HF + CKD, HF + anemia, CKD + anemia), or for CRAS overall, were statistically significant, indicating no synergistic interaction.

**Table 3:** Evaluation of interactions among CRAS components.

|  | ACD or HF admission |  |
| --- | --- | --- |
|  | HR (95% CI) | P-value |
| Age | 1.04 (1.02–1.06) | <0.001 |
| Body mass index | 0.95 (0.92–0.99) | 0.015 |
| Systolic blood pressure | 0.99 (0.99–1.00) | 0.034 |
| Male | 1.20 (0.94–1.54) | 0.147 |
| MR severity | 1.15 (0.82–1.58) | 0.412 |
| HF | 2.29 (1.11–5.32) | 0.022 |
| CKD | 1.97 (0.52–6.29) | 0.252 |
| Anemia | 2.39 (1.00–6.04) | 0.052 |
| HF + CKD | 0.50 (0.14–2.07) | 0.352 |
| HF + anemia | 0.56 (0.20–1.49) | 0.232 |
| CKD + anemia | 0.85 (0.22–3.77) | 0.812 |
| CRAS | 2.32 (0.46–10.56) | 0.297 |

## Discussion

In this national, real-world cohort from the REVEAL-AFMR registry, the main findings were as follows: (1) approximately 25% of patients with AFMR had CRAS; and (2) CRAS independently predicted higher rates of future cardiovascular events, including ACD and HF hospitalization. Interaction analysis showed that HF and anemia contributed to poor cardiovascular outcomes and had an additive effect on overall prognosis, whereas no statistical synergy among CRAS components was observed, indicating that CRAS represents a clinically cumulative risk burden rather than a multiplicative interaction. These findings highlight the combined impact of HF, CKD, and anemia in patients with AFMR and emphasize the need for comprehensive risk assessment and integrated management.

### Mechanism of CRAS and the association of AFMR

Silverberg et al. first described the term CRAS in 2002 as the coexistence of chronic HF, CKD, and anemia (4). Each component reflects systemic organ dysfunction and exacerbates the others through interrelated pathophysiological mechanisms (5). First, HF reduces renal perfusion and triggers neurohormonal activation, thereby impairing kidney function. Second, kidney dysfunction activates the renin–angiotensin–aldosterone system, leading to fluid retention, myocardial hypertrophy, necrosis, and fibrosis, thereby worsening HF. Kidney dysfunction also suppresses erythropoietin production, leading to decreased erythrocyte production in the bone marrow, uremic toxin accumulation, and anemia. Finally, anemia further compromises oxygen delivery to peripheral tissues, activates the sympathetic nervous system, and increases cardiac workload, thereby accelerating left ventricular remodeling and HF progression. This vicious cycle creates synergistic deterioration and is strongly associated with adverse clinical outcomes. In the context of AFMR, CRAS may exert deleterious effects on a specific hemodynamic status. Elevated LA pressure, chronic venous congestion, and reduced forward cardiac output may have detrimental effects on renal function and anemia. In our study, the prevalence of overlapping conditions was as follows: HF and CKD in 351 (35.6%) patients, HF and anemia in 437 (44.3%), CKD and anemia in 312 (31.6%), and CRAS in 258 (26.2%). HF, CKD, and anemia are often observed in patients with AFMR.

### CRAS and prognosis

CRAS predicts poorer outcomes across various cardiovascular settings. In both acute and chronic HF, regardless of left ventricular ejection fraction, CRAS is associated with higher rates of hospitalization and mortality, with a reported prevalence of 19%–62% and mortality as high as 51% (12–16). CRAS is also associated with poor prognosis in patients at risk for HF (17), those with acute myocardial infarction (18), and individuals with cancer (19). In MR, renal dysfunction (20) and anemia (21) have been associated with poor prognosis after transcatheter mitral valve repair; however, the impact of CRAS on the prognosis of patients with MR remains unclear.

To our knowledge, this is the first study to investigate the prognostic value of CRAS in patients with AFMR. We found that each component of CRAS—HF, CKD, and anemia—was individually associated with adverse outcomes in patients with AFMR. The coexistence of these conditions in CRAS exerted an additional adverse effect on prognosis, worsening clinical outcomes beyond those associated with any single component. This association between CRAS and adverse clinical outcomes remained consistent regardless of the severity of MR or treatment strategy. Therefore, CRAS represents a key risk factor in patients with AFMR and warrants comprehensive evaluation and management.

### Clinical implications

Our findings highlight the potential benefits of targeting CRAS components, particularly HF and anemia, in the management of patients with AFMR. In this cohort, CRAS predicted adverse outcomes, irrespective of MR severity or treatment strategy. These findings underscore the importance of early recognition and comprehensive CRAS management to improve outcomes, regardless of surgical, transcatheter, or medical therapy.

Although data on the population with AFMR are limited, recent advances in HF management offer potential avenues for improving outcomes in patients with CRAS. Sodium-glucose cotransporter 2 inhibitors increase hematocrit levels and alleviate anemia, in addition to improving prognosis in patients with HF and CKD (22). Similarly, timely rhythm control or structural interventions for AFMR may interrupt the cycle of hemodynamic deterioration in patients with CRAS. The management of anemia and iron deficiency in patients with HF has garnered increasing attention in recent years, and treatment with intravenous iron has improved clinical outcomes (23–25). Conversely, the role of erythropoiesis-stimulating agents in renal anemia remains controversial (26–29), and evidence for hypoxia-inducible factor prolyl hydroxylase (HIF-PH) inhibitors is still emerging; early studies, however, suggest potential benefits (30).

The current clinical guidelines do not provide specific evidence-based recommendations for the management of patients with AFMR and CRAS. Nevertheless, early identification of and intervention for CRAS—including volume optimization, anemia correction, and renal protection—should form the cornerstone of treatment for this high-risk group. Future prospective studies are warranted to clarify optimal management strategies and improve outcomes.

### Study limitations

The present study has some limitations. First, this was an observational registry study involving a heterogeneous patient population. No standardized treatment protocols were used across the participating centers, which may have affected patient outcomes. The impact of specific anemia treatments, including iron supplementation, erythropoiesis-stimulating agents, and HIF-PH inhibitors, was not assessed. Second, longitudinal changes in renal function, hemoglobin levels, MR severity, and treatment response were not investigated. Data were evaluated at the time of enrollment; however, subsequent changes may have influenced the prognosis. Third, the etiologies of renal dysfunction and anemia were not determined. Levels of iron, total iron-binding capacity, transferrin, transferrin saturation, reticulocytes, vitamin B12, red cell distribution width, and hematocrit were unavailable. Prognostic differences based on the underlying causes of anemia and CKD may also be important and should be investigated in future studies.

Despite these limitations, CRAS remained a significant risk factor for future cardiovascular events in patients with AFMR.

## Conclusion

CRAS is common in patients with AFMR and is independently associated with a high risk of future cardiovascular events. These results emphasize the need for early recognition and comprehensive management of CRAS in this population. Further prospective investigations are warranted to determine whether targeted interventions against CRAS components can improve outcomes in patients with AFMR.

## Clinical Perspective

### What is new?

This is the first study to demonstrate that cardio-renal anaemia syndrome (CRAS) is an independent predictor of poor outcomes in patients with atrial functional mitral regurgitation (AFMR). CRAS was common in patients with AFMR, affecting approximately 25% of this population. The coexistence of heart failrue, chronic kidney disease, and anemia demonstrated an additive adverse effect on prognosis. Among these components, heart failure and anemia were particularly strong drivers of poor prognosis. These associations remained consistent regardless of mitral regurgitation severity or treatment strategy.

### What are the clinical implications?

In patients with AFMR, identification of CRAS may be valuable for risk stratification and prognosis prediction. Targeted treatment of CRAS components—especially heart failure and anemia—may play a key role in improving prognosis among patients with AFMR. Further prospective trials are needed to evaluate whether tailored therapies—such as sodium-glucose cotransporter 2 inhibitors, hypoxia-inducible factor prolyl hydroxylase inhibitors, or intravenous iron supplementation—can improve prognosis in patients with AFMR and CRAS.

## Data Availability

The data that support the findings of this study are available from the corresponding author upon reasonable request.

## Acknowledgements

We thank the study investigators for their contributions. We thank Editage (www.editage.jp) for English language editing.

